# Association between adipokines and glycemia in children under age 8: the PROGRESS cohort of Mexico

**DOI:** 10.64898/2026.01.20.26344469

**Authors:** Yara S. Beyh, Haotian Wu, K.M. Venkat Narayan, Lisa R. Staimez, Usha Ramakrishnan, Manoj K. Bhasin, Maricruz Tolentino-Dolores, Guadalupe Estrada-Gutierrez, Maayan Yitshak-sade, Robert Wright, Martha M. Téllez-Rojo, Andrea A. Baccarelli

**Affiliations:** Emory Global Diabetes Research Center, Emory University, Atlanta, Ga, USA; Department of Environmental Health Sciences, Mailman School of Public Health, Columbia University, New York, NY, USA; Hubert Department of Global Health, Rollins School of Public Health, Emory University, Atlanta, GA, USA; Department of Biomedical Informatics, Emory University, Atlanta, GA, USA; Research Branch, National Institute of Perinatology, Mexico City, Mexico; Department of environmental medicine and public health, The Icahn School of Medicine at Mount Sinai, New York, NY, USA; Center for Nutrition and Health Research, National Institute of Public Health, Mor. Mexico; Department of Environmental Health, T. H. Chan School of Public Health, Harvard University, Boston, MA

## Abstract

**Background:** The role of adipokines in childhood glycemia is poorly understood. We investigate the longitudinal association between adipokines and glycemia in a cohort of children in Mexico City.

**Methods:** Children from the **Programming Research in Obesity, Growth, Environment, and Social Stressors** (PROGRESS) cohort (948 children, 52% male) were followed longitudinally from birth. Leptin, adiponectin, glucose, and HbA_1c_ were measured at four, six, and eight years, and fasting insulin at eight years. Adiponectin to leptin ratio (ALR) and HOMA2 indices were computed. Longitudinal associations were examined by linear mixed models and cross-sectional associations were examined by multivariable linear regression. All models were adjusted for maternal and child covariates.

**Findings:** Between ages four and eight years, average levels of leptin increased from 3·2 to 10·8 ug/mL; adiponectin dropped from 15·7 to 13·7 ng/mL; and ALR dropped from 9·1 to 3·1 ug/ng. Longitudinally, across timepoints four, six, and eight years after birth, there was no association between adipokines and glycemia. However, the cross-sectional analysis at age 8 years found an association between leptin and insulin (1·0, 95% CI: 1·0; 1·1), HOMA2-B (1·0, 95% CI: 1·0; 1·0), HOMA2-IR (1·0, 95% CI: 1·0; 1·1), and HOMA2-S (0·9, 95% CI: 0·9; 0·9).

**Interpretation:** Further investigation is needed to understand the role of adipokines in the development of T2DM in children and the factors that may alter adipokine metabolism.

## I. Introduction

Once thought to be an adult disease, Type 2 Diabetes Mellitus (T2DM) in the pediatric population is an aggressive illness characterized by a rapid decline in pancreatic beta cell function, high treatment failure, and more aggressive complications (e.g., cardiovascular complications, etc(1–5)). T2DM prevalence in children has increased over the last two decades in tandem with an increase in child obesity(1,2,5), especially among ethnic minority groups(4,6,7). For instance, the prevalence of T2DM in Mexico City among youth increased from 20·2% in 2013 to 33·0% in 2018, which accounts for a 12·8% significant increase(8). Yet, there exists significant gaps in our understanding regarding the development and treatment of pediatric T2DM.

Adipose tissues (adipocytes) have an endocrine function through releasing a number of adipokines, some of which are anti-inflammatory (e.g. adiponectin), while others are pro-inflammatory (e.g., leptin)(9–13). The latter stimulate inflammation and insulin resistance, whereas the former have a protective role. Thus, any imbalance between the two families of adipokines is associated with diseases such as diabetes and are believed to cause alterations in insulin sensitivity increasing metabolic risks(12,13). It has been suggested that elevated leptin or reduced adiponectin levels are connected to insulin resistance (IR) in T2DM in adults, linking them through obesity(10,11,14,15). Given the opposing effects of leptin and adiponectin, some researchers have postulated that the two adipokines interact with each other in the modulation of T2DM risk, with adiponectin having the stronger effect(10,16), thus, suggesting the use of the ratio between the two adipokines as a useful index for IR.

Unfortunately, the studies examining the associations between adipokines and glycemia have largely focused on adolescents and adults, leaving scant data in children in the prepubertal ages. We investigated the contribution of adipokines to the development of glycemic dysregulation in childhood by examining the longitudinal association between adipokines and glycemia using multiple timepoint data from a cohort of children in Mexico City.

## II. Materials and methods

### a. Study participants

In this study, we used data from the Programming Research in Obesity, Growth and Social Stress (PROGRESS) Study, a prospective birth cohort based in Mexico City. Working with the Mexican Social Security Institute (IMSS) prenatal clinics, we enrolled women in the 2^nd^ trimester of pregnancy between 2007 and 2010 followed them till delivery (n=948). Children were followed after birth in six months intervals till age of two years and every two years thereafter. Study details have been previously described(17,18). Women receiving prenatal care from IMSS were eligible if they were at least 18 years of age; gestational age less than 20 weeks; having a singleton pregnancy; planning to reside in Mexico City for the next three years; free of heart or kidney disease; not using steroids or anti-epilepsy drugs; do not consume alcohol on a daily basis; and have access to a telephone. Participants signed written consent forms and children provided assents after age six. After enrollment, study visits occurred in 2^nd^ and 3^rd^ trimester and delivery for mothers. Mothers and children were seen at one month, six months, 12 months, 18 months, 2-3 years, 4-5 years, 6-7 years, and 8-9 years postpartum. All study protocols were approved by the Institutional Review Board (IRB) at Columbia University Human Research Protection Office (protocol #AAAR0689), Icahn School of Medicine at Mount Sinai (protocol #12-00751), and the Mexican National Institute of Public Health (protocol #560).

### b. Outcome and exposure measurement

The exposure measurements were leptin, adiponectin, and adiponectin-to-leptin ratio (ALR). The main outcome measurements were glucose and glycated hemoglobin (HbA1_c_). At the 4-5 years blood samples were not collecting following fasting given their very young age. At the 6-7 years, and 8-9 years of age samples were collected in the morning following an overnight fast. Trained phlebotomists collected and prepared the child’s venous blood samples per standard protocol. Ultrasensitive ELISA measured adiponectin (R&D Systems, Minneapolis, MN) and leptin (ALPCO Diagnostics Inc. Salem, NH) at all visits. All measurements had less than 2% coefficients of variations between replicate samples. Missing values from the adiponectin measurements (N=2) and from the leptin measurements (N=3) occurred due to technical errors. ALR was computed by dividing the value of adiponectin by that of leptin to be used in the analysis. HbA1_c_ was measured in whole blood at all visits by a particle enhanced immunoturbidimetric test (Response 910 Analyzer). Plasma glucose was measured at all visits by enzymatic photometric assays on a Response 910 automated analyzer (DiaSys Diagnostic Systems, Holzheim, Germany).

The secondary outcome measures were insulin and insulin resistance (HOMA2-B, HOMA2-S, and HOMA2-IR). For this, we used plasma insulin, which was measured at visit 8-9 years by a solid-phase, enzyme-labeled chemiluminescent immunoassay, on an Immulite 1000 analyzer (Siemens, Germany). Using available glucose and insulin measurements at visit 8-9 years, HOMA-2 indices were computed with the HOMA-2 calculator developed by the Diabetes Trial Units at the Oxford Center for Diabetes, Endocrinology, and Metabolism (Oxford University, UK) to estimate steady state beta cell function (%B), insulin sensitivity (%S), and insulin resistance (%IR).

### c. Covariates

Prenatal covariate information was obtained from baseline questionnaires. Child covariates were obtained from baseline questionnaire and the four year follow-up visit. An index for socioeconomic status (SES) was calculated based on the Mexican Association of Research and Public Opinion Agencies (AMAI) version 13×6 that was collapsed into a relative three-level index of lower, medium, and higher(19). We included both maternal and child covariates based on previous studies(20–23) and the needs for the current analysis. Maternal covariates included: age at enrollment, pre-pregnancy BMI (Kg/m2) using predicted mother’s pre-pregnancy weight, gestational weight gain (Kg), parity (primiparous or multiparous), educational level (less than high school, high school, or more than high school), SES which was collapsed into three categories (low, medium, or high) reflecting the study population and not necessarily the population in Mexico City, smoking status during pregnancy (Yes or No), alcohol consumption during pregnancy (Yes or No), and breastfeeding status at 1-month post-partum (never breastfed, attempted, non-exclusively, or exclusively). Child covariates included: gestational age (weeks), sex (female or male), sedentary level (minutes/day), physical activity level (minutes/day), and age at each visit.

### d. Statistical analysis

Given the longitudinal nature of the study, we assumed that missing values for certain variables were at random. The rest of the measurements from these individuals were included in our analyses. The variable with the highest missingness adiponectin at the four year follow-up (n=461), and the variable with the least missingness was leptin at the six year follow-up (n=523). To deal with the missing data, we ran 50 multiple imputations. The number of iterations was based on the highest percentage of missingness with a slight inflation to keep the length of the confidence interval close to that of a complete case analysis(24–26).

We examined the simultaneous associations between the exposures (leptin, adiponectin, and ALR) and glucose or HbA1_c_, using linear mixed models with random intercepts for each child to account for repeated measurement design. We adjusted the models for maternal age at enrollment, maternal pre-pregnancy BMI, gestational weight gain, parity, educational level, SES, smoking status during pregnancy, alcohol consumption during pregnancy, and breastfeeding status at one month post-partum, gestational age, sex, sedentary and physical activity levels at four years of age, and child’s age at the time of the follow-up visit.

Additionally, we used multivariable linear regression to estimate the associations between our exposures and each of insulin, insulin sensitivity, HOMA2-IR, HOMA2-S, and HOMA2-B measured at eight years of age, which were all log2 transformed prior to analysis to improve model fit. For these transformed outcomes, effect estimates were expressed as the percent relative change of the outcome for every one unit increase in exposure. We built models of individual exposures and outcomes and mutually adjusted models with all biomarkers to examine the differences in the results. Child sex was examined as an effect measure modifier by running a stratified analysis.

All analyses were conducted using SAS 9.4.

## III. Results

### a. Study participants characteristics

At time of enrollment, maternal age was on average 27·7 years (SD = 5·5) and mothers had an average predicted pre-pregnancy BMI(27) of 26·3 Kg/m^2^ (SD = 4·2). Most of the participants came from a lower-medium SES background (n=842, 88·8%), had at least a previous pregnancy (n=593, 62·6%), had an educational status of high school or less (n=719, 75·8%), reported not smoking during pregnancy (n=602, 99·0%), the majority reported consuming alcohol during the 2^nd^ trimester of pregnancy (n=850, 89·7%) compared to the 3^rd^ trimester (n=25, 3·1%). At a follow-up visit, most women reported not exclusively breastfeeding at one-month post-partum (n=469, 60·83%) and had an average gestational weight gain of 7·5 Kg (SD = 3·6). Maternal characteristics are reported in **Table 1**.

**Table 1.** Maternal characteristics at enrollment (N=948)

| Characteristics | Means (SD) |
| --- | --- |
| Age at enrollment (years) | 27.66 (5.49) |
| Pre-pregnancy BMI (Kg/m<sup>2</sup>)† | 26.29 (4.16) |
| Gestational weight gain (Kg) | 7.5 (3.59) |
| Breastfeeding status at 1-month post-partum N [%] |  |
| Never breastfed | 52 (6.74) |
| Attempted | 47 (6.10) |
| Non-exclusive | 469 (60.83) |
| Exclusive | 203 (26.33) |
| Missing | 177 |
| Alcohol consumption during pregnancy N [%]* |  |
| Yes | 850 (89.66) |
| No | 98 (10.34) |
| Smoking status during pregnancy N [%]* |  |
| Yes | 6 (0.99) |
| No | 602 (99.01) |
| Missing | 340 |
| Educational level N [%] |  |
| Less than high school | 385 (40.61) |
| High school | 334 (35.23) |
| More than high school | 229 (24.16) |
| Socio-economic status of the household‡ N [%]** |  |
| Lower | 486 (51.27) |
| Medium | 356 (37.55) |
| Higher | 106 (11.18) |
| Parity N [%]*** |  |
| Yes | 355 (37.45) |
| No | 593 (62.55) |

\*Asked during the second trimester.
\*\*Calculated based on AMAI criteria and collapsed into 3 categories (lower, medium, higher) for modelling.
\*\*\* Collapsed into primiparous and non-primiparous for modelling.
†Computed using predicted maternal pre-pregnancy weight.
‡Collapsed into three categories from six originally, hence, reflective of the study population and not the general population of Mexico City.

Children born to recruited women had a quasi-equal split between males and females (52·2% and 47·5% respectively) with average gestational age of 38·2 weeks (SD = 1·9) and an average BMI z-score of -0·9 (SD = 1·3) at birth. At the 8-9 years follow-up visit, the mean adiponectin, leptin, ALR, fasting glucose, HbA1_c_, and fasting insulin were 13·7 ng/mL (SD = 15·9), 10·8 ug/mL (SD = 10·8), 3.1 ug/ng (SD = 1·4), 4·8 mmol/L (SD = 0·6), 4·91% (SD = 4·9), and 66·9 pmol/L (SD = 57·5), respectively. The computed ALR, HOMA2-IR, HOMA2-B, and HOMA2-S values were 3·1 ug/ng (SD = 1·4), 1·2 (SD = 1·0), 115·9 (SD = 66·1), and 157·3 (SD = 172·9), respectively. Child characteristics are reported in **Table 2**.

**Table 2.**
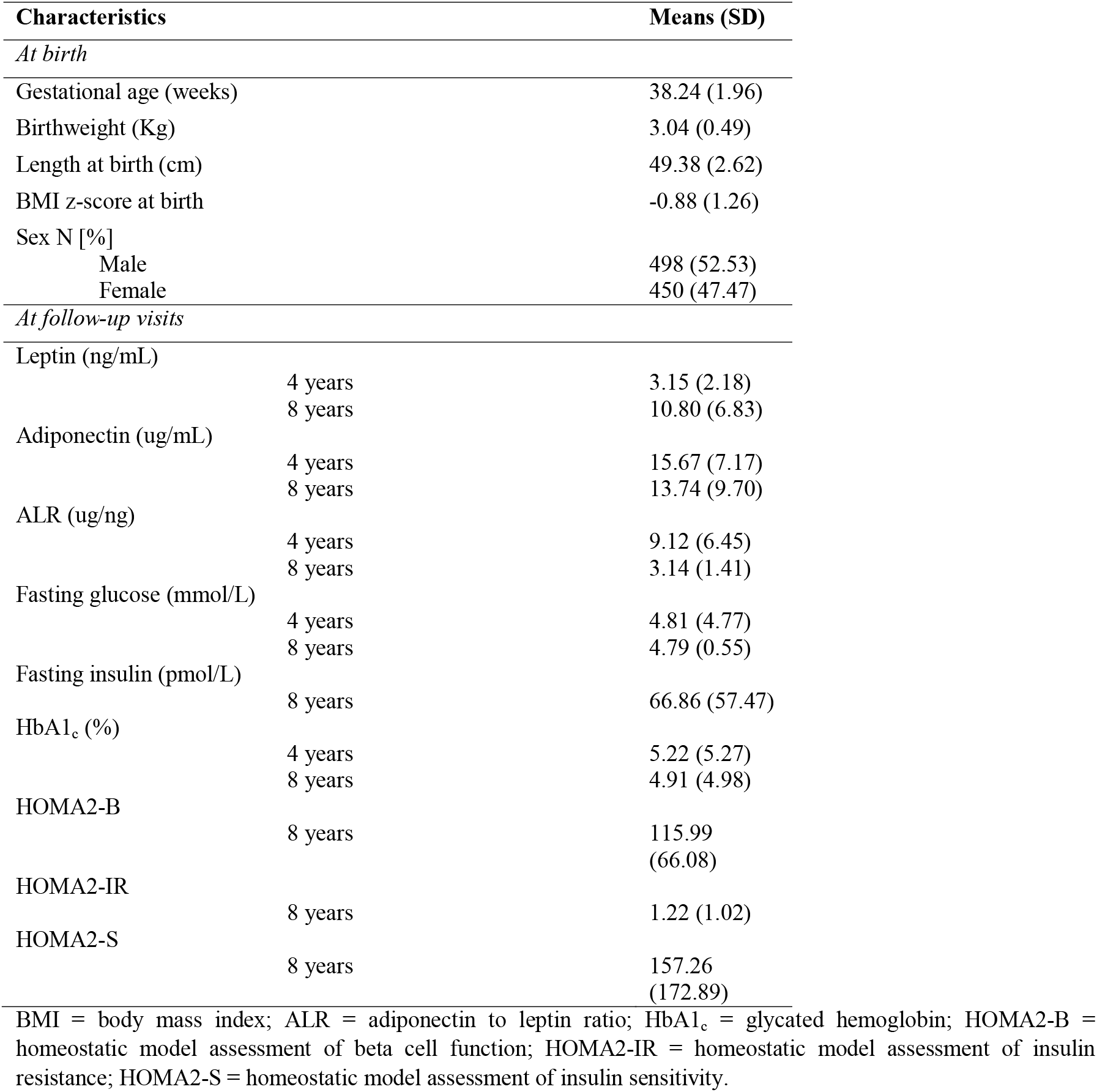
Child characteristics at birth and 8 years of age (N=948) Characteristics Means (SD)

### b. Main association of child adipokines and glycemic measures

#### i. Longitudinal analysis: Glucose and HbA1_c_

Follow-up visits at six and eight years of age are referred to as Time 2 and Time 3, respectively, in **Table 3**. We investigated the change in outcomes (glucose and HbA1_c_) over time in relation to the exposures of interest (leptin, adiponectin, and ALR). The interaction term between adipokine and time-point reflects whether the adipokine had any influence at that specific age. The analysis showed that only ALR influenced glucose concentrations at age six with an absolute change of -0·03 (95% CI: -0·05, -0·01). Whereas there was no effect for any of the adipokines on the absolute change in HbA1_c_ at any time point. Associations were similar for the unadjusted model and the model adjusted for all the maternal and child covariates mentioned above.

**Table 3.** Associations of adipokines at 96 months of age with glucose and HbA1_c_ from 48 months to 96 months of age.

|  | Percent change | 95% confidence interval |
| --- | --- | --- |
| **Glucose** |  |  |
| Leptin | 0.01 | -0.03; 0.04 |
| Leptin*Time 2 | 0.00 | -0.04; 0.04 |
| Leptin*Time 3 | 0.01 | -0.03; 0.04 |
| Adiponectin | -0.01 | -0.02; 0.01 |
| Adiponectin*Time 2 | -0.01 | -0.03; 0.01 |
| Adiponectin*Time 3 | 0.00 | -0.02; 0.02 |
| ALR | 0.00 | -0.01; 0.01 |
| ALR*Time 2 | -0.03* | -0.05; -0.01 |
| ALR*Time 3 | -0.01 | -0.03; 0.02 |
| **HbA1<sub>c</sub>** |  |  |
| Leptin | 0.00 | -0.02; 0.02 |
| Leptin*Time 2 | 0.01 | -0.02; 0.03 |
| Leptin*Time 3 | 0.01 | -0.02; 0.03 |
| Adiponectin | 0.01 | -0.01; 0.01 |
| Adiponectin*Time 2 | 0.00 | -0.01; 0.01 |
| Adiponectin*Time 3 | 0.00 | -0.01; 0.01 |
| ALR | -0.01 | -0.01; 0.00 |
| ALR*Time 2 | 0.01 | -0.01; 0.02 |
| ALR*Time 3 | 0.01 | -0.02; 0.02 |

ALR = adiponectin to leptin ratio; HbA1<sub>c</sub> = glycated hemoglobin; Time 2 = 72 months of age; Time 3 = 96 months of age. The reference point is 48 months of age.
Model adjusted for mother's age, gestational age, gestational weight gain, mother's BMI (using predicted weight), maternal smoking status during pregnancy, maternal alcohol consumption during pregnancy, breastfeeding status at 1-month post-partum, socio-economic status of the household, maternal educational level, parity, child sex, child sedentary activity level at 48 months of age, child's physical activity level at 48 months of age, and child's age at the follow-up visit.

#### ii. Cross-sectional analysis: Insulin, HOMA2-S, HOMA2-IR, and HOMA2- B

Results from each of the outcomes are presented in **Table 4**. Multivariable linear model analysis examining the associations between adipokines (leptin, adiponectin, ALR) and each of insulin, insulin sensitivity, HOMA2-S, HOMA2-IR, and HOMA2-B showed a significant association between leptin and each of the outcomes at 96 months of age. For every one unit increase in standard international unit of leptin, there was a 3% increase in relative change in insulin (95% CI: 1·0; 1·1); 3% decrease in relative change in insulin sensitivity (95% CI: 0·9; 0·9); 2% increase in relative change in HOMA2-B (95% CI: 1·0; 1·04); 3% increase in relative change in HOMA2-IR (95% CI: 1·00; 1·07); and 3% decrease in relative change in HOMA2-S (95% CI: 0·94; 0·99). Adiponectin and ALR did not show any significant associations with any of the outcomes. Associations were similar for the unadjusted model and the model adjusted for all the maternal and child covariates mentioned above.

**Table 4.**
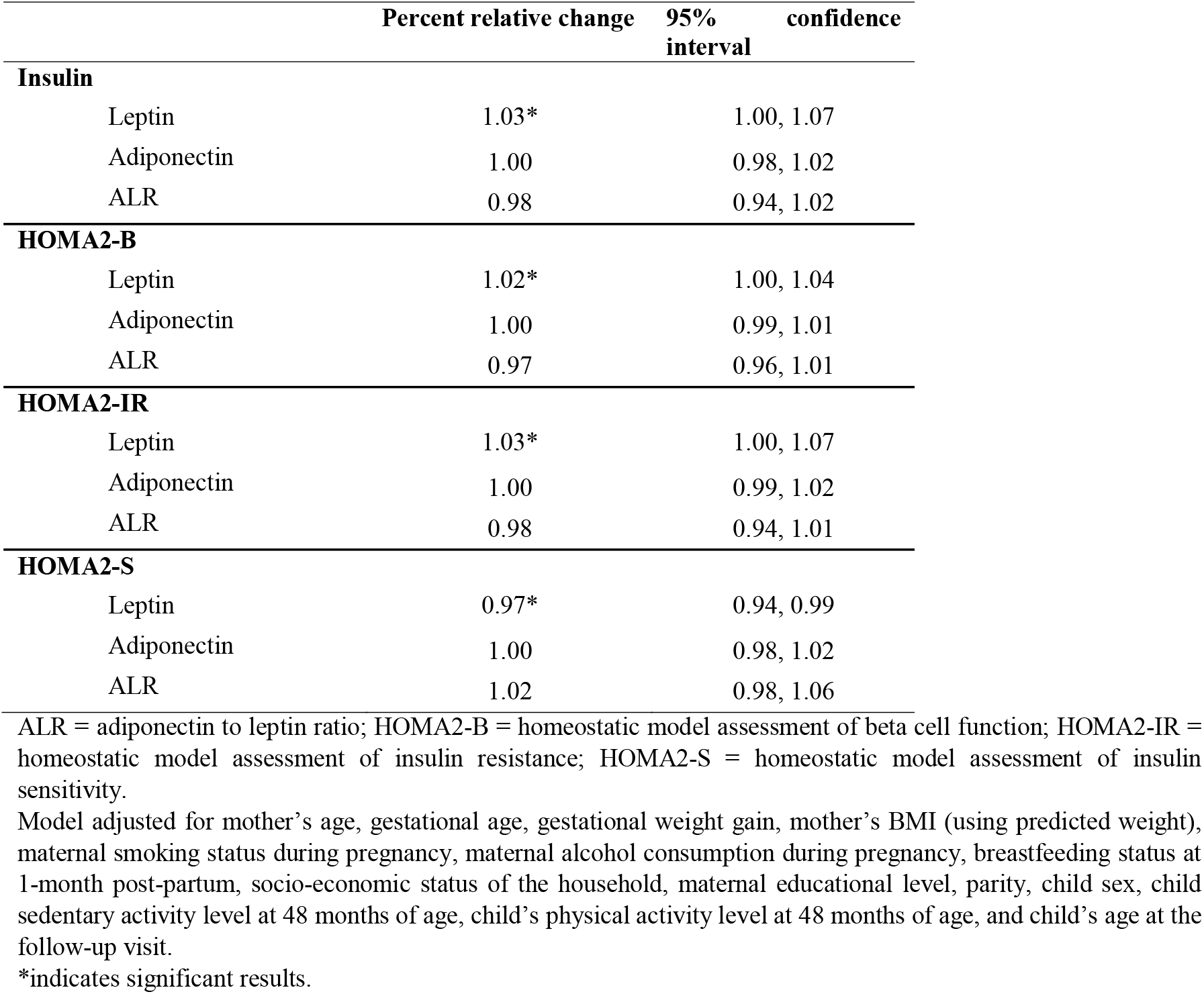
Associations between adipokines and glycaemia measures at 96 months of age.

### c. Effect modification by child sex

We investigated whether child sex would modify the associations of adipokines with glycemic measures. Sex stratified models were adjusted for both maternal and child covariates described previously. First, we examined whether child sex would influence the associations of adipokines with glucose and HbA1_c_ over time. We took the values at the first measurement (age four) as the reference and reported the percent change of glucose and HbA1_c_ at six and eight years per unit increase of the adipokines. The results shown in **Figure 1** (A-Leptin, B-Adiponectin, and C-ALR) show similar percent changes between males and females at six and eight years of age, hence, the non-existence of a statically significant difference between the genders. Albeit, a noticeable trend is worth highlighting, which is the increased change at age six followed by the decreased change at eight years of age, for both glucose and HbA1_c_, while glucose showed a decrease at both time points.

Further, the stratified analysis returned no statistically significant difference between males and females in the association between adipokines and insulin measures. The results are presented in **Figure 2** (A-Leptin, B-Adiponectin, and C-ALR). The percent relative change in insulin, insulin resistance, HOMA2-IR, HOMA2-B, and HOMA2-S were similar between males and females with every increase in one standard international unit in the exposures. In some instances, females had a slightly higher relative change in the associations compared to males, but it was not statistically significant.

## IV. Discussion

Obesity and diabetes are becoming more prevalent in adults and adolescents and are accompanied by a disruption in adipokines levels. We proposed to determine whether adipokine disruption is also seen in pre-pubertal children. In our study population followed form birth until the age of 8 years, adipokines followed a similar pattern. In our longitudinal analysis examining the association between adipokines, and glycemic measures (glucose and HbA1_c_) we did not find evidence of adipokine disruption. Those results contrast with the prior studies on association between adipokines and glycemia shown in other studies conducted in adolescents and adults(28–30). Because these are pre-pubertal children, one possible explanation would be the little change observed in glucose and HbA1c values between the ages of four and eight years, as described in **Table 2**. Hence, our study population had an improvement in their HbA1_c_ levels with unchanged fasting glucose, which would make an association between adipokines and glycemia difficult to observe.

As for the multivariable linear analysis, it showed that, at age eight years, there is an association between leptin and insulin (**Table 4**). Our results regarding leptin were consistent with previous studies conducted in adults and adolescents, where leptin was positively associated with IR and higher concentrations were found in people with diabetes(31–34), as well as obese children between the ages of six and 18 years(29). Further, the null association we found between adiponectin and glycemia measures was previously observed in a study conducted on children between the ages of 12-16 years, whereas leptin was found to be associated with IR and beta cell dysfunction(35). Other studies in adults and adolescents found that adiponectin is protective against IR and diabetes, with decreased concentrations in obesity and diabetes(11,31,33,36). Other studies found associations between ALR and IR measures in normoglycemic individuals, which is not the case in our study. Further, researchers recommended the use of ALR as an index of IR rather than using HOMA-IR or glucose measures, especially as fasting glucose levels are usually elevated in people with diabetes(29,37,38). Additionally, the conclusion from a study examining association of ALR and IR in children (12-16 years) was that ALR is a marker of IR across obesity classes I and II, but not in normal weight and obesity class III individuals(29).

Additionally, children weight gain is based more on fat-free mass than fat mass in pre-pubertal stages, whereas adipokines are proportionate to the fat mass in the body(39,40). Thus, the levels of adipokines in children might have not been elevated enough to detect associations between adipokines and glycemia at this age.

### Proposed mechanisms

Leptin and adiponectin are two adipokines that play roles in insulin resistance and glucose homeostasis. Under normal conditions, glucose-lowering effects of leptin are mediated through various organs. In the muscles, leptin improves insulin sensitivity by decreasing intramyocellular lipid levels and activating cAMP-activated kinase (AMPK); whereas in the liver, leptin has the capacity to reduce intracellular hepatic triacylglycerol levels(13). However, elevated concentrations of leptin have the capacity to downregulate insulin secretion from pancreatic beta cells under elevated glucose concentrations in circulation; in addition to inhibiting insulin gene expression(11,13,41). Specifically, leptin inhibits glucose-stimulated insulin secretion through the cAMPK-A or protein kinase C (PKC) pathways. Leptin has a direct effect on insulin through the hyper-polarization of beta cell membranes by opening the ATP-sensitive potassium channels (KATP), which in turn reduces the intracellular calcium involved in releasing insulin from the vesicles of beta cells(11,41). This effect is still not fully understood, but can be explained by three possible mechanisms of leptin: 1) stimulation of phosphodiesterase 3B that hydrolyzes cAMP to AMP, as well as the activity of phosphoinositide-3-kinase, hence reducing the level of cAMP needed for closing KATP channels(11,42); 2) stimulated formation of long-chain acyl-CoA esters known to open KATP channels(11,43); and 3) reduced transport of glucose into beta cells, hence lowering the concentration of ATP, which in low ratio to adenosine diphosphate can open KATP channels(11,44).

As for adiponectin, under normal circulating concentrations, its action is known to improve insulin sensitivity through the activation of AMPK in both the liver and the skeletal muscles(11,41,45). In fact, this is achieved through adiponectin binding to its receptors AdipoR1 and AdipoR2, in the muscles and liver, respectively(11,31,41). This action leads to the increased glucose uptake in the muscles and reduced gluconeogenesis in the liver(11,31,41,45). Other proposed insulin-sensitizing mechanisms include reduction of triacylglycerol content in tissues; upregulation of insulin signaling; and activation of PPARα(11,13).

This study is one of the very few that investigated the longitudinal association between adipokines and glycemia between the ages of four and eight years, in addition to insulin resistance, in pre-pubertal children from a Mexican ethnicity. Hence, even the most modest results could contribute to the slim knowledge we have about the role of adipokines in diabetes in the pediatric population. It is also important to note that the study had a large sample size with an almost equal distribution between males and females. Additionally, ample data was collected at baseline and at follow-up visits, from both mothers and children, to be used as covariates in the analysis phase.

Our study had also its limitations. For instance, insulin was measured only at 8 years of age, hence measures of the homeostatic model assessment (HOMA2-IR, HOMA2-S, and HOMA2-B) were computed only at one time-point, preventing us from including insulin resistance measures as part of the longitudinal analysis. Further, in an ideal study, we would have included the disposition index (DI) and the insulinogenic index (ISI) as they provide more information about the glycemic status of our sample population, however, we did not have fasting and 30-mintue glucose and insulin assays, which are needed to compute DI And ISI. Additionally, certain variables had observations missing completely at random. We addressed this issue by implementing multiple imputations on our data rather than excluding participants who missed some visits. Finally, data was collected from women and their offspring in Mexico City and had access to healthcare; hence, excluding people living in rural areas or lack medical attention.

## V. Conclusion

We found an association between leptin and each of insulin, HOMA2-B, HOMA2-S, and HOMA2-IR at the age of eight years. However, we were unable to detect a significant association between the other adipokines and insulin measures, nor longitudinally with glycemia measures. Further, our results did not show any differences between males and females at this age.

This study was an initial investigation into the associations between adipokines and glycemia, as well as insulin resistance, in pre-pubertal children from Mexican ethnicity. Based on our findings, there is strong suggestions that adipokines might be playing a role in the development of diabetes at younger ages. Hence, we suggest future work be conducted on other ethnicities who suffer from a large incidence rate of diabetes (e.g., Asian Indians, etc.). This will provide us with a better understanding of the metabolic role of the adipokines in the development and progression of diabetes in children.

## Data Availability

All data produced in the present study are available upon reasonable request to the authors.

## Abbreviations

ALR: adiponectin-to-leptin ratio
AMAI: Mexican Association of Research and Public Opinion Agencies
AMPK: cAMP-activated kinase
%B: beta cell function
BMI: body mass index
CDC: Centers for Disease Control and Prevention
CI: confidence interval
CR: concentration ratio
FCS: fully conditional specification
HbA1_c_: Hemoglobin A1_c_
HOMA: homeostatic model assessment
%IR: insulin resistance
IMSS: Mexican Social Security Institute
IRB: Institutional Review Board
KATP: ATP-sensitive potassium channels
PKC: protein kinase C
PROGRESS: Programming Research in Obesity, Growth and Social Stress
%S: insulin sensitivity
SES: socioeconomic status
SOCS: suppressor of cytokine signaling
T2DM: type 2 diabetes

## Authorship contribution statement

**Yara S. Beyh**: Methodology, Software, Formal analysis, Interpretation, Writing – original draft, Writing – review and editing, Visualization. **Haotian Wu**: Methodology, Interpretation. **Lisa R. Staimez**: Supervision, Writing – review and editing. **Usha Ramakrishnan**: Supervision, Writing – review and editing. **Manoj K. Bhasin**: Supervision, Writing – review and editing. **K.M. Venkat Narayan**: Supervision, Writing – review and editing. **Andrea A. Baccarelli**: Funding acquisition, Project administration, Supervision, Writing – review and editing. **Maricruz Tolentino-Dolores**: Lab tests, Writing – review and editing. **Guadalupe Estrada-Gutierrez**: Lab tests, Writing – review and editing. **Maayan Yitshak-Sade**: Writing – review and editing. **Robert Wright**: Funding acquisition, Writing – review and editing. **Martha M. Téllez-Rojo:** Funding acquisition, Writing – review and editing.

## Acknowledgments

We gratefully acknowledge all the members of the PROGRESS team for their tireless efforts in maintaining the study cohort. We would also like to thank the study participants and PROGRESS staff for making this work possible.

This work has been presented as a poster presentation at the 83^rd^ American Diabetes Association Scientific Session “Diabetes 2023” (https://diabetesjournals.org/diabetes/article/72/Supplement_1/1097-P/150765/1097-P-Association-between-Adipokines-and-Glycemia)

## Disclaimer

The findings in this article reflect the opinions of the authors and do not necessarily represent the opinion of the Centers for Disease Control and Prevention. The use of trade names is for identification only and does not imply the endorsement by the CDC, the Public Health Service, or the US Department of Health and Human Services.

